# Home Food Procurement Impacts Food Security and Diet Quality during COVID-19

**DOI:** 10.1101/2021.02.02.21251017

**Authors:** Meredith T. Niles, Kristen Brassard Wirkkala, Emily H. Belarmino, Farryl Bertmann

## Abstract

**Background:** Home food procurement (HFP) (i.e. gardening, fishing, foraging, hunting, backyard livestock and canning) have historically been important ways that people obtain food. Recently, some HFP activities have grown (e.g. gardening), while other activities (e.g. hunting) have become less common in the United States. Anecdotally, COVID-19 has sparked an increase in HFP evidenced by increased hunting licenses and shortages in seeds and canning supplies. HFP may have positive benefits for food security and diet quality, though research beyond gardening is especially limited in high-income countries.

**Methods:** We examine HFP activities before and since the COVID-19 pandemic, and their relationship to food security and dietary quality using multivariable logit models and matching analysis with a statewide representative survey (n=600) of residents of Vermont, United States.

**Results:** We find 29% of respondent households classified as food insecure since COVID-19, and more prevalence among those experiencing a negative job change since COVID-19, households earning less than $50,000 annually, Hispanic and multi-race respondents. Forty-two percent of respondents engaged in HFP activities; the majority of those gardened, and more than half pursued HFP activities more intensely than before the pandemic. HFP was more common among food insecure households, who were more likely to fish, forage, hunt and have backyard livestock. Respondents who were food insecure, Black, Indigenous, People of Color and/or Hispanic, those with a negative job disruption, and larger households all had greater odds of increased intensity of HFP since COVID-19. HFP were significantly associated with eating greater amounts of fruits and vegetables, especially if gardening and canning, while respondents hunting or having backyard livestock were significantly more likely to have higher red meat intake.

**Conclusion:** Overall, these results suggest that HFP activities have increased since the start of the COVID-19 pandemic, and may be an important safety net for food insecure households, and provide diet quality impacts. Long-term, HFP activities may have important food security and diet quality impacts, as well as conservation implications, which should be more thoroughly explored. Regardless, the increased interest and intensity of HFP demonstrates multiple opportunities for educational and outreach efforts.

## Background

The COVID-19 pandemic has highlighted the uncertainty and fragility of food security and food access globally. In the United States, unemployment rates reached unprecedented levels at their height in April 2020, causing concerns among many Americans about how to access affordable and high-quality food [1]. Existing evidence suggests that home food procurement (i.e. backyard livestock, fishing, foraging, gardening, hunting, and canning, hereafter referred to as HFP) may offer opportunities to improve food security and diet quality (e.g. [2, 3]). HFP activities have varying levels of participation in recent decades. While homesteading [4] and backyard livestock, especially chickens, have become more fashionable in recent years [5], hunting has been declining for decades [6, 7]. However, since the COVID-19 pandemic began, there have been a number of stories from popular media outlets in the United States discussing a comeback of “victory gardens” in response to the pandemic [8, 9], increased interest and demand for hunting and fishing [10], and a shortage of canning supplies [11]. As well, previous research has found that depictions of wild food foraging in the media change in times of economic hardship from being discussed as more of a luxury to being conceptualized as a way to provide for basic needs [12]. Public discussion and interest around HFP practices seem to be shifting with COVID-19, but who is participating and what relationship do these activities have to food security and dietary outcomes? This study explores changes in HFP since the onset of the COVID-19 pandemic, and its relationship to food security and diet quality outcomes during the pandemic in a high-income country context.

The potential for HFP to improve food security and dietary outcomes has links to other challenging times, including in historical moments such as World War 2. At that time, planting “victory gardens” were patriotic acts to grow local food amidst disrupted supply chains [13]. It is estimated that 40% of the nation’s fruits and vegetables were produced via victory gardens during the war, demonstrating the potential for HFP to address food security challenges. But the current COVID-19 context has created new difficulties and significant increases in food insecurity in many countries, including the United States (e.g.[14, 15]). Nevertheless, existing evidence suggests that HFP may positively affect both food security and dietary quality outcomes in high-income countries through multiple pathways.

First, evidence suggests that growing your own food contributes directly toward food availability and access. Taylor & Lovell (2015) found that, while gardeners did not grow enough to sustain their families, ⅓ grew a substantial quantity and were self-sufficient in providing some items for a certain period of time during the growing season and almost all of these households said they always had enough to eat. Corrigan’s (2011)[16] interviews of five gardeners in Baltimore found that most perceived that they saved money from their gardens and that it allowed them to grow quality, fresh produce that otherwise may not have been accessible. They also found that many gardeners canned or froze their excess produce, allowing them access to these foods into the winter. These results may also translate beyond gardening to other food procurement practices, although research is even more limited in these areas. Smith et al. (2019) [17] found that participants from one reservation who participated in the Food Distribution Program on Indian Reservations who also so hunted, fishes, or foraged were more food secure than those who did not. Additionally, those who engaged in more than one practice were more secure than those who did only one. A survey of Canadian Inuit also found that households with an active hunter were more food secure than those without an active hunter [18]. Cooke et al. (2018) [19] found that many anglers in the United States often consume what they catch, with an average of 4,700 grams of edible fish provided through fishing annually, even if their original motivation for fishing is recreation. As well, African American anglers are more likely to consider fishing important for providing food [20] and more likely to keep fish that they have caught [21], though these studies did not examine food security outcomes. This direct food procurement may also lead to cost savings realized by not purchasing food, which enable money to be available for the purchase of other foods, or for other financial priorities.

Realized cost savings from HFP may be another factor linking HFP to better food security outcomes. Perceived cost savings does appear to be a common motivation for those producing their own food [17, 22] and there have been a number of studies suggesting that this may in fact be the case [2, 3, 23, 24]. Home gardeners in San Jose reported that cost savings of gardening allowed them to eat produce they otherwise would not have had access to [25]. However, many studies looking at cost savings were analyzing the results of nonprofit programs in which gardeners were supported with resources to help set up their gardens, and therefore had a smaller up-front investment, which could have impacts on food security outcomes. Csortan et al. (2020) [26] found that 65% of gardeners would break even on garden investments in five years or less and then start saving money. In such a case, gardening would not be a sufficient means for achieving food security in the short-term in response to an economic crisis. They also found that the number of years of gardening experience appeared to have a positive impact on productivity and resource efficiency, leading to additional concerns for new gardeners [26].

In addition to the potential for cost savings and increased food security, HFP may lead to a higher quality, more diverse diet, including one that may be more culturally appropriate [27, 28]. Growing one’s own produce is linked to increased fruit and vegetable intake [3, 25, 29–31]. Hunting, fishing, and foraging may also lead to a more nutritious and diverse diet; for example, 80% of people surveyed on a native reservation said that hunting, fishing, and foraging made their diets more diverse and 72% said these practices improved the quality of their diet [17]. Stark et al. (2019) [32] found that wild edible greens were abundant in three low-income neighborhoods in California, and offered potential nutrient density comparable to some common nutritious vegetables, such as kale. Some research suggests that growing one’s own food may also lead to improved nutritional knowledge [33, 34] and changes in eating habits for the long-term [24, 34, 35]. This may also be true of children, who are more likely to try vegetables when they garden [36].

Prior research suggests that partaking in HFP strategies may lead to an increase in food security and diet quality outcomes, but the current research is limited, especially as it pertains to the impact of hunting, fishing, and foraging practices in high-income countries. Further, COVID-19 has changed the way many people work, live, and shop, potentially providing opportunities or new barriers to HFP and new challenges for food security and high-quality diets. Emerging evidence indicates that dietary quality has decreased during the COVID-19 pandemic in many places (e.g. [37, 38], offering potential opportunities for HFP to counter such trends. Existing evidence of HFP activities since the start of the COVID-19 pandemic is limited, though our previous work found about half of respondents reported producing, foraging, hunting, or canning last year and nearly one third were engaging in those activities at the time of the survey [39] (Belarmino et al., 2020). Chenarides et al. (2020) [40] examined urban gardening before and during the COVID-19 pandemic, finding lower participation in community gardens as compared to at-home gardens. Constant et al. (2020) found having a garden/terrace positively associated with unhealthy behaviors during the COVID-19 lockdown in France, primarily a reduction in physical activity. Finally, though a few commentaries have discussed the potential benefits of home gardens during COVID-19 (e.g.[41, 42]), to our knowledge, no population-based studies have comprehensively assessed HFP activities during the pandemic and its relationship to food security and diet quality outcomes. This study aims to fill this gap by surveying a representative sample of people in Vermont to understand their HFP strategies, change in activity during the first five months of the COVID-19 pandemic, and the relationship of HFP to food security, and diet quality. In a predominantly rural state such as Vermont, these concerns are especially pressing, as rural areas are estimated to have 50% higher rates of food insecurity than urban areas [43].

## Methods

### Survey development and sampling strategy

The data were collected using a survey instrument developed initially in March 2020 [44], in collaboration with other researchers as part of the National Food Access and COVID research Team (NFACT) [45]. The survey was further refined [46], with the later forming the basis for this data collection. The survey measures multiple components of food access, food security, dietary quality, home food procurement, COVID-19 experiences and food assistance program participation as well as individual and household sociodemographics. Institutional Review Board approval was obtained from The University of Vermont (IRB protocol 00000873) prior to any data collection. The survey utilizes validated measurements when possible (Table 1), and was also validated prior to release of Version 1 in Vermont with 25 eligible (18 and over) respondents using Cronbach alpha and factor analysis [14]. All question sets obtained an internal validity of alpha > 0.70 [47, 48].

**Table 1.**
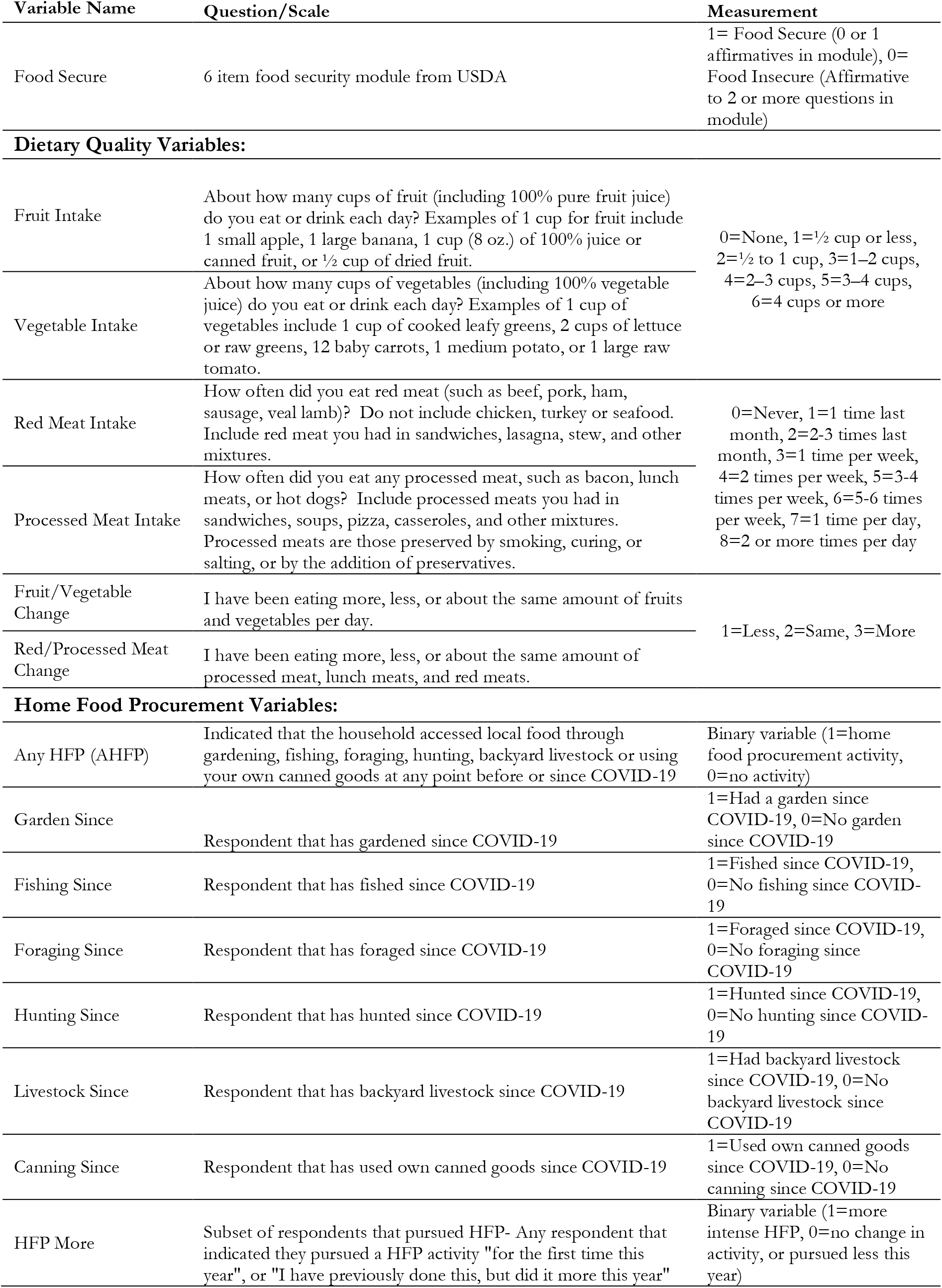
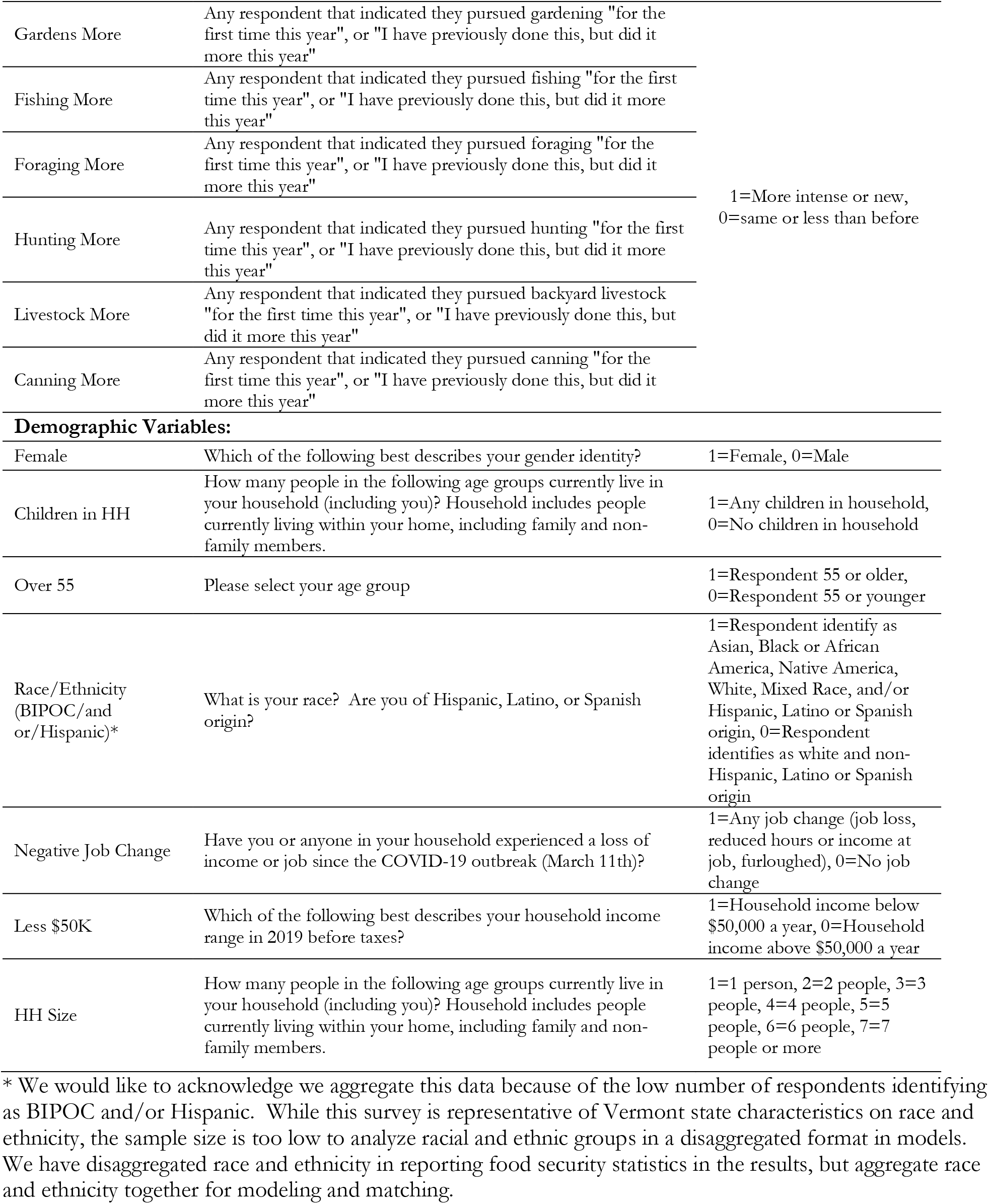
Complete list of variables, questions and measurement utilized in this analysis.

Participants were recruited through an online survey administered by Qualitrics (Provo, UT), using a general population sample representative to the state of Vermont with respect to income, race and ethnicity. This sample was achieved by matching sample recruitment quotas to the income, race (White, Black or African American, American Indian and Alaska Native, Asian, Native Hawaiian or Other Pacific Islander, and Two or more races), and ethnicity (Hispanic, non-Hispanic) population profile of Vermont in the American Community Survey [49]. A total of 600 people ages 18 and over responded to the survey, representing a margin of error (95% confidence level) for this segment of the Vermont population of +/- 4% [50]. The survey was administered in August and September 2020.

### Variables of Interest

We explore three self-reported dependent variables in this analysis (Table 1). First, food security status, as measured through the US Department of Agriculture 6-item short-form food security module [51] where food insecurity is classified as answering affirmatively to two or more out of six questions. This was modified to ask respondents about food security since COVID-19 (approximately five months at the time of the survey) rather than the traditional 12-month period. Second, current fruit and vegetable intake was measured through the National Cancer Institute’s 2-item screener [52], which was modified to apply to the last month and some example foods were removed to shorten it. Current red and processed meat intake was measured using two questions from the Dietary Screener Questionnaire in the National Health and Nutrition Examination Survey (NHANES) 2009-10. Finally, we developed new questions to measure perceived change in fruit/vegetable, red meat, and processed meat consumption since the onset of the COVID-19 pandemic. Independent variables included multiple questions related to previous and current HFP, specific HFP activities, and changes in HFP activities during the COVID-19 pandemic, as well as several household and individual-level demographics (Table 1).

### Statistical analysis

We utilize a series of logistical regression models, reporting with odds ratios to examine how demographic factors correlate with any home food procurement (AHFP), and the different HFP strategies. We use chi-square tests to examine food security and diet quality changes since the start of the COVID-19 pandemic as it relates to AHFP, specific HFP activities, and intensity of HFP. We use one-way analysis of variance (ANOVA) to examine diet quality intake at the time of the survey as it relates to AHFP, specific HFP activities, and intensity of HFP. Then, to examine how AHFP, intensity of HFP, and specific HFP activities relate to both food security outcomes and dietary quality, we use nearest neighbors matching techniques.

Matching techniques are useful with observational data to estimate causal effects of treated and control groups, aiming to balance the distribution of covariates across treated and control groups [53]. Here we explore how AHFP, intensity of HFP, or specific HFP activities are “treatments” on food security and diet quality, using demographic factors as matching covariates across groups. We use six demographic covariates in our matching analysis: female, children in household (HH), race/ethnicity (Black, Indigenous, People of Color (BIPOC)/and or Hispanic), negative job change, household income less than $50,000 (less $50k), and HH size (Table 1), which are likely to be associated with the treatment and outcome [54, 55]. Matching techniques also require defining a distance (measure of similarity between the individuals). We use a nearest neighbor matching approach with a Mahalanobis distance, which accounts for covariance among variables, and is documented to work well with fewer than eight covariates [56, 57]. For each treated individual, nearest neighbor matching selects a control individual with the smallest distance from that individual. For example, if we are exploring AHFP, the technique would have people who did and did not engage in AHFP as “treatment” and control groups, and then match a treatment and control respondent together based on similar demographic covariates included in the analysis (e.g. household size and job change status). In all our models we use nearest neighbor matching with five matches per observation, meaning each observation was matched with five closest other observations within the control and treatment groups.

## Results

### Respondent Characteristics

Table 2 details the specific respondent characteristics, which reflect the demographic composition of the Vermont population for the gender, race, and income distribution. Overall, 67.3% of the respondents were female (std. dev= 0.47), and 30.2% of respondents had children in the household (std. dev= 0.46). Forty-four percent of respondents were age 55 years or older. Reflecting the racial/ethnic profile of Vermont, 8.3% of respondents identified as BIPOC and/or Hispanic ethnicity (std. dev= 0.28). More than 46% of respondents lived in a household that had experienced a negative job change during the first five months of the COVID-19 pandemic (job loss, loss of income or hours from job, or furlough) (std. dev=0.50). Household size was on average 2.57 (std. dev= 1.34), with 60.2% of households 2 or fewer people.

**Table 2.**

| Characteristic |  | Respondents (N=600) |
| --- | --- | --- |
| Age - no. (%) | 18-34 | 153 (25.5) |
|  | 35-54 | 182 (30.3) |
|  | 55+ | 263 (43.8) |
| Children in household - no. (%) | Yes | 178 (30.2) |
|  | No | 415 (70.0) |
| Gender - no. (%) | Female | 404 (67.3) |
|  | Male | 190 (31.7) |
|  | Transgender/Non-binary/Self-Described | 6 (1.0) |
| BIPOC - Race - no. (%) | White | 559 (93.2) |
|  | Two or more races | 22 (3.7) |
|  | American Indian or Alaska Native | 5 (0.8) |
|  | Asian | 4 (0.7) |
|  | Black or African American | 9 (1.5) |
| BIPOC - Ethnicity - no. (%) | Not Hispanic or Latino | 583 (97.2) |
|  | Hispanic or Latino | 17 (2.8) |
| 2019 Household Income - no. (%) | Less than \$10,000 per year | 39 (6.5) |
| | \$10,000-\$24,999 | 81 (13.5) |
| | \$25,000-\$49,999 | 141 (23.5) |
| | \$50,000-\$74,999 | 110 (18.3) |
| | \$75,000 - \$99,999 | 77 (12.8) |
| | \$100,000 or more | 145 (24.1) |
| Job change during the COVID-19 pandemic - no. (%) | Lost job | 149 (24.8) |
|  | Reduced hours or income | 208 (34.7) |
|  | Furloughed | 122 (20.3) |
|  | Any change | 270 (46.2) |
|  | No changes | 314 (53.8) |
| Household Size - no. (%) | 1 to 2 | 357 (60.2) |
|  | 3 to 5 | 211 (35.6) |
|  | 6 or more | 25 (4.2) |

### Descriptive Statistics of Key Variables

Among all respondents, 42.1% (n=250) engaged in AHFP activity during the first six months of the COVID-19 pandemic, with the greatest number of respondents gardening (34.7%), followed by canning (23.5%) and fishing (10.2%) (Figure 1). Among only respondents who engaged in AHFP, 51.8% (n=128) did at least one HFP activity more intensely since the COVID-19 pandemic began or for the first time this year, with the greatest increase in intensity of activity among backyard livestock (52%, n=26), gardening (45.3%, n=106), and foraging (44.9%, n=31).

**Figure 1.**
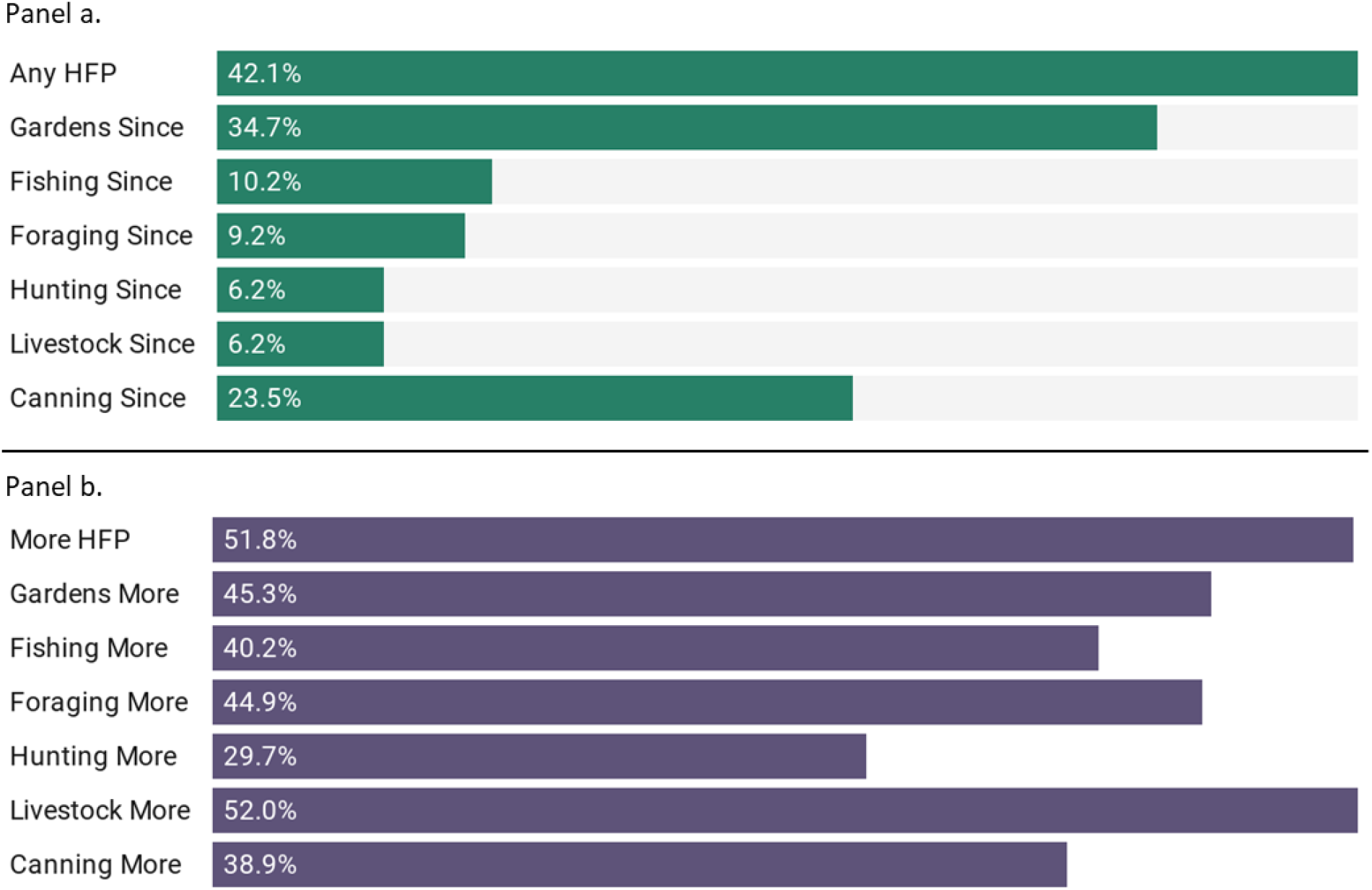
a) Percent of respondents engaging in any HFP, and specific HFP activities since COVID-19. Percentages include all respondents (n=600). b) Among respondents who engaged in any HFP (n=250), percent of those that increased intensity or did a new HFP activity since COVID-19.

On average, respondents self-reported they ate between 1-2 cups cumulatively of fruit (mean=2.20) and vegetables (mean=2.74) per day, though 11% and 5% of respondents ate no fruit or vegetables respectively daily. Respondents self-reported they ate red meat (mean=3.34) and processed meat (mean=3.15) about one time per week, with 10% each indicating they never eat red or processed meat. Nearly one in four (23.3%) respondents indicated eating less fruits and vegetables during the pandemic as compared to before, 65.5% reported eating the same as before COVID-19, and 11.2% reported eating more. Changes in red and/or processed meat consumption were also indicated by about one-third of respondents, with 25.9% eating less red and/or processed meat since the start of the COVID-19 pandemic, and 7.9% eating more.

### Demographics of Food Security

Among our dependent variables, 29.0% (n=169) of respondent households were classified as food insecure at some point since COVID-19. To assess the relationship of our demographic controls on food security, we ran a logit model (Supplementary Table 1). Respondents 55 and over were at higher odds of food security (OR=2.52, p=0.001), while households experiencing a negative job disruption (OR=0.47, p=0.001), and those earning less than $50K annually (OR=0.134, p<0.001), were at reduced odds of food security. Disaggregating race and ethnicity demonstrates higher rates of food insecurity among multiple race respondents (33.3%) and Hispanic respondents (50%) as compared to non-multi-race respondents and non-Hispanic respondents (p<0.10). Black respondents showed the highest overall rate of food insecurity (50%), however, these results are not statistically significant with a chi-square test, likely because of our low sample size (Supplementary Table 2).

### Demographics of Home Food Procurement

Using a multivariate logit model, we examine how demographics correlate with different aspects of AHFP. We find that households experiencing a negative job change have 1.81 greater odds (p=0.001) of any AHFP (Table 3), while households earning less than $50,000 annually were at reduced odds (0.694, p=0.043) of AHFP. Among those that did AHFP, we find that multiple demographic factors are correlated with increased intensity of HFP during the pandemic. Specially, BIPOC /Hispanic respondents (OR=3.58, p=0.026), households experiencing a negative job change (OR=1.89, p=0.026), and larger households (OR=1.48, p=0.021) were at significantly greater odds of increased intensity of HFP while respondents over 55 were at significantly reduced odds of increasing intensity during the pandemic (OR= 0.49, p= 0.029) (Table 4).

**Table 3.** Multivariate logit model predicting any home food procurement activities with demographics.

| Variable | Odds Ratio | Std. Error | P= | 95% Confidence Interval |  |
| --- | --- | --- | --- | --- | --- |
| Female | 1.058 | 0.201 | 0.768 | 0.728 | 1.537 |
| Children in HH | 1.158 | 0.299 | 0.569 | 0.699 | 1.920 |
| Over 55 | 1.122 | 0.234 | 0.583 | 0.745 | 1.689 |
| BIPOC/Hispanic | 1.081 | 0.341 | 0.804 | 0.583 | 2.005 |
| Negative Job Change | 1.812 | 0.325 | 0.001 | 1.274 | 2.576 |
| Less \$50K | 0.694 | 0.125 | 0.043 | 0.487 | 0.989 |
| HH Size | 0.908 | 0.080 | 0.271 | 0.764 | 1.079 |

**Table 4.**
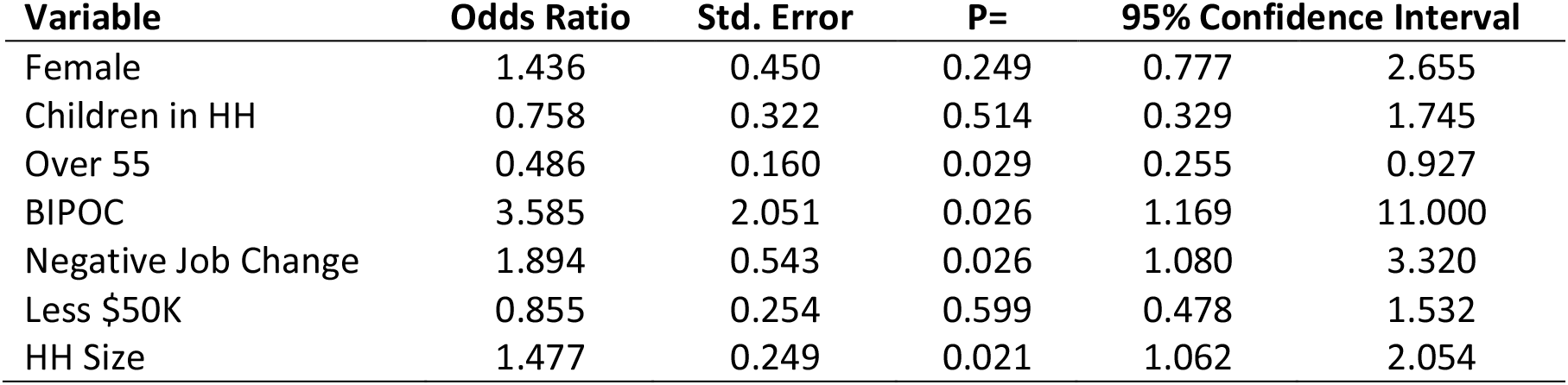
Multivariate logit model predicting increased intensity of home food procurement since COVID-19 with demographics.

Multivariate logistical regression models predicting the specific types of all six HFP activities by demographics found multiple significant factors. Respondents with a negative job change were at increased odds of gardening (OR=1.43, p=0.055), while households making less than $50,000 annually were at reduced odds (0.63, p=0.014). Respondents over 55 were at reduced odds of fishing since the start of the COVID-19 pandemic (OR=0.50, p=0.051), while respondents over 55 (OR=1.85, p=0.097), and respondents with a negative job change (OR=2.13, p=0.014) were at increased odds. Women were at reduced odds of hunting during the pandemic (OR=0.46, p=0.034, while households with children were at increased odds (OR=2.26, p=0.094). Respondents over 55 were at reduced odds of having backyard livestock during the pandemic (OR=0.16, p=0.001), while households with a negative job change were at increased odds of canning (OR=1.45, p=0.067) and households making less than $50,000 annually were at reduced odds of canning (0.703, p=0.091) (Supplementary Tables 3-8).

### Home Food Procurement and Food Security

AHFP was more common among food insecure households (47.3%) as compared to food secure households (39.2%) (p=0.076). Overall, food insecure respondents were significantly more likely to be fishing (p=0.005), foraging (p=0.003), hunting (p<0.001), canning (p=0.019), and have backyard livestock (p=0.008) during the COVID-19 pandemic (Figure 2).

**Figure 2.**
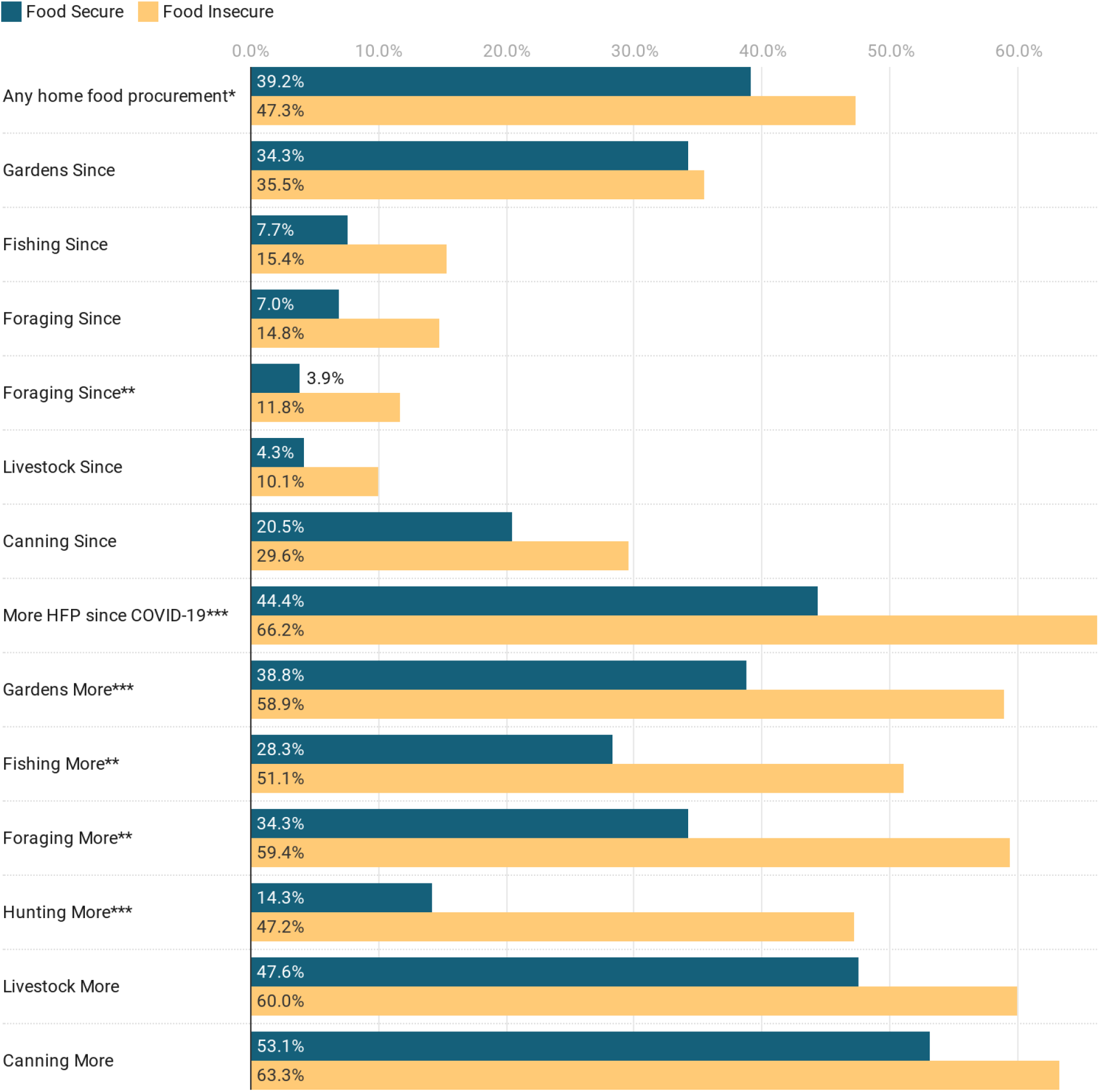
Percent of food secure and food insecure households engaging in various types of HFP activities and intensity since COVID-19. (*=p<0.10, **=p<0.05, ***=p<0.01, Supplementary Table 9). Questions about any HFP, and specific HFP since COVID-19 include all respondents. Questions about increased HFP activity are asked only of respondents engaging in HFP (n=250).

Using a matching approach, to examine the effect of AHFP on household food security we find a weak negative association between AHFP and food security (b=-0.070, p=0.059), while controlling for gender, children in the household, negative job change, income, race/ethnicity, and household size. Exploring the effect of specific HFP activities during the pandemic on food security outcomes, we find that fishing (b=-0.133, p=0.038), foraging (b=-0.173, p=0.025), hunting (b=-0.264, p=0.021), and canning (b=-0.100, p=0.017) are all negatively associated with food security (Table 5).

**Table 5.**
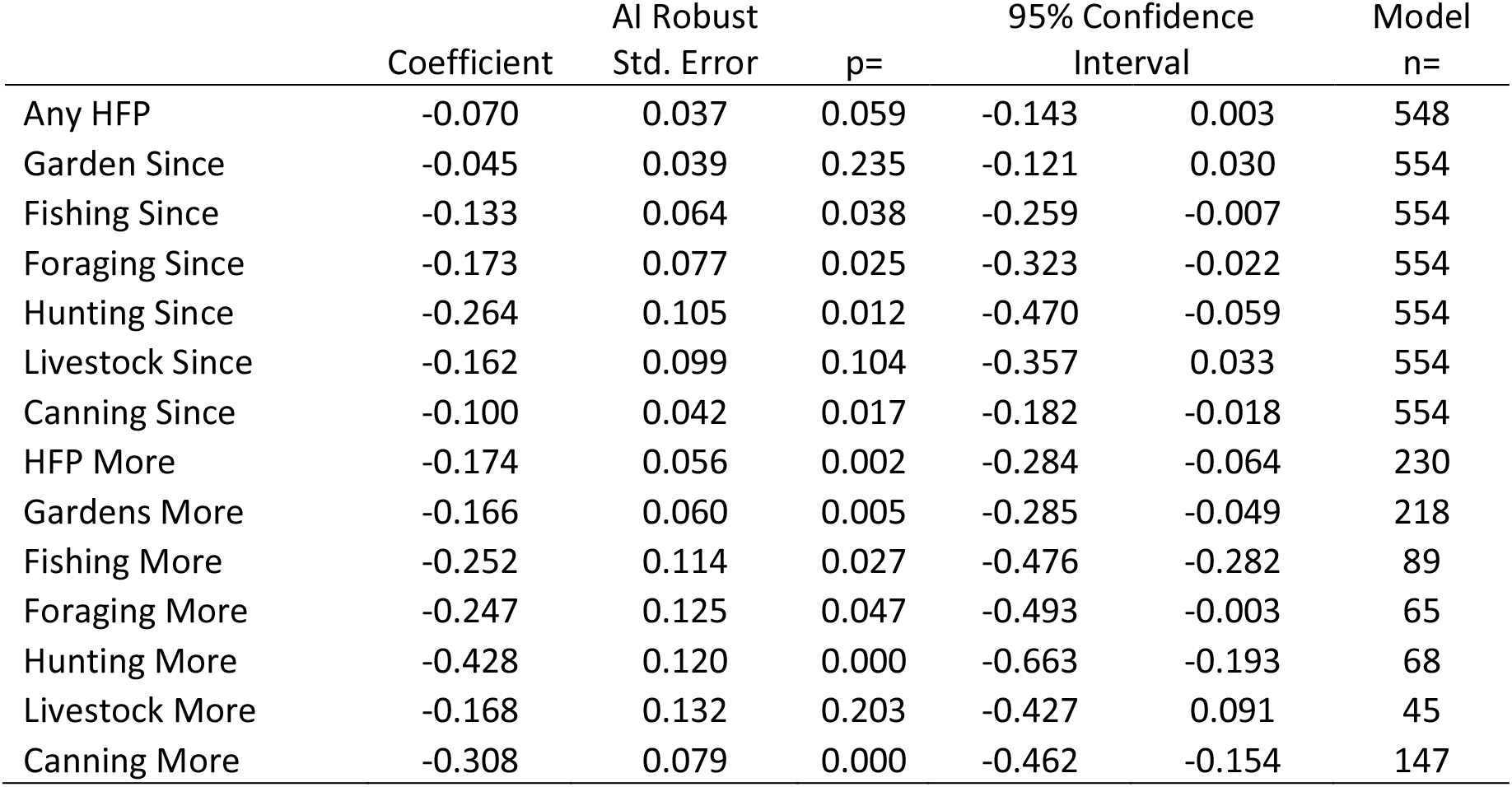
Food security outcomes as related to HFP using nearest neighbors matching analysis. Each row indicates a separate matching analysis, where the HFP variable was used as a “treatment” while using six demographic controls (Female, Children in HH, BIPOC, Negative Job Change, Less $50k, HH size) to conduct the matching.

We also find through chi-square analysis, significant associations between food security and intensity of HFP since the COVID-19 pandemic began, with 66.2% of food insecure households increasing intensity of HFP since the COVID-19 pandemic began, compared to 44.4% of food secure households (p=0.002). Food insecure households were also more likely to be gardening (p=0.005), fishing (p=0.025), foraging (p=0.040), and hunting (p=0.003) more intensely than before the COVID-19 pandemic as compared to food secure households (Figure 2). These results are confirmed by matching analysis, controlling for demographics (p<0.05, Table 5).

### Home Food Procurement and Diet Quality

We use ANOVA to examine the current dietary quality at the time of the survey as it relates to AHFP, specific HFP activities and intensity of HFP. Respondents engaging in AHFP were significantly more likely to eat greater amounts of fruits (mean 2.42 compared to 2.06, p=0.001) and vegetables (mean 3.04 compared to 2.55, p<0.001) (Figure 3). We find no significant differences between AHFP and intake of red meat (mean=3.45 compared to 3.29, p=0.284) or processed meat (mean=3.06 compared to 3.21, p=0.309). Using matching techniques, with demographic controls, we examine current fruit, vegetable, red meat and processed meat intake as it relates to AHFP, increased HFP, and relevant specific HFP activities (i.e. gardening, foraging and canning for fruit and vegetable intake and fishing, hunting and backyard livestock for red and processed meat intake). We find the “treatment” of AHFP to have a significant and positive relationship to higher fruit (b=0.386, p=0.001) and vegetable intake (b=0.526, p<0.001). Furthermore, we find that gardening and canning since the COVID-19 pandemic began have significant effects on higher current intake of fruits (gardening b= 0.329, p=0.006; canning b=0.240, p=0.071) and vegetables (gardening b=0.541,p <0.001; canning b= 0.511, p<0.001) (Supplementary Tables 10 and 11). We find no significant effect of AHFP or increased intensity of HFP on current red meat or processed meat intake; however, we do find a significant effect of hunting (b=0.527, p=0.032) and backyard livestock (b=0.794, p=0.001) since the start of the COVID-19 pandemic on current red meat intake, with higher red meat intake among those engaged with hunting and/or backyard livestock production (Supplementary Tables 14 and 15).

**Figure 3.**
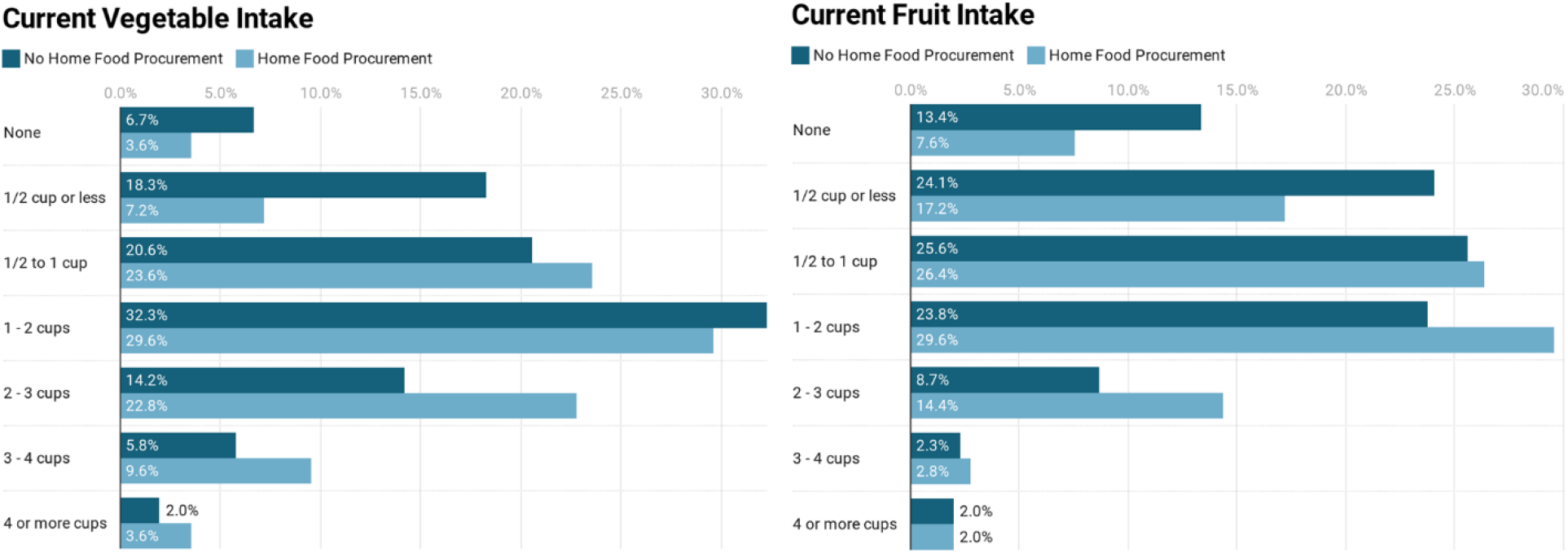
Current self-reported vegetable and fruit intake among respondents engaged or not in HFP. On average, respondents engaged in HFP are significantly more likely to be eating more fruit and vegetables (p≤0.001).

We use chi-square tests to examine the change in dietary quality outcomes since the COVID-19 pandemic began as it relates to AHFP, specific HFP activities and intensity of HFP. We find households engaging in AHFP have a higher proportion of respondents with increased fruit and vegetable intake (14.8% compared to 8.4%, p=0.051), while those engaging in AHFP have both a higher proportion of respondents eating less red meat (29.2% compared to 23.3%) and a slightly higher proportion of respondents eating more red meat (9.6% compared to 6.7%, p=0.079). Using matching techniques, with demographic controls, we examine change in fruit/vegetable and red/processed meat intake as it relates to AHFP, increased HFP, and relevant specific HFP activities (i.e. gardening, foraging and canning for fruit and vegetable intake and fishing, hunting and backyard livestock for red meat intake). We find no significant effects of AHFP, increased intensity, or specific HFP activities on change in fruit and vegetable intake since the start of the COVID-19 pandemic. We do find that increased intensity of hunting is negatively associated with meat intake since the COVID-19 pandemic began, suggesting that those hunting more, or for the first time during the pandemic are eating less red meat.

## Discussion

Overall, we find a significant increase in HFP since the beginning of the COVID-19 pandemic, evidence that has been documented in the popular media, but not yet widely shown through peer-reviewed literature. Those engaging in AHFP were more likely to be in low-income households and those with negative job changes, and increased intensity was more likely among those with negative job changes, BIPOC/Hispanic respondents, and larger households. Importantly, food insecure households were more likely to be using AHFP, and especially more likely to have increased intensity of HFP during the pandemic. While we find that nearly 25% are eating less fruits and vegetables since before the onset of the COVID-19 pandemic, we also find that AHFP is positively associated with higher fruit and vegetable intake. These results were especially prevalent among gardening and canning households, while red meat intake was higher among households hunting and having backyard livestock.

These results have several important implications. First, it suggests that food insecure households engage in HFP as a potential coping mechanism for food insecurity, and this appears to have been especially true during the first US growing season during the COVID-19 pandemic. This is further corroborated by the results that low-income households and those with negative job changes were also more likely to be engaging in HFP and increasing the intensity of their engagement. While food insecure households are overall more likely to engage in AHFP (both before and since the start of the COVID-19 pandemic), more than 2/3 of food insecure households engaged more intensely in HFP or for the first time during the first five months of the pandemic. It is also important to note that a higher percentage of food insecure households are engaging more in non-gardening HFP activities (e.g. hunting, fishing, foraging) during the pandemic. One recent change in Vermont law provides free hunting and fishing licenses to certified members of state-recognized Native American tribes, which may have influenced some Native Americans to pursue these activities for the first time, or more intensely since COVID-19. Coupled together, these results provide important evidence about the reliance on HFP during a pandemic, and as a “safety net” for many potential households engaging in these activities for the first time or more intensely than before.

Our results are counter to some of the existing research that demonstrates that households using HFP are more food secure than those not using HFP [17], though the existing research on this topic is limited. There are several potential explanations for these different findings. First, the existing research in a Western context generally has had small sample sizes (e.g. [16]), and often focused on specific populations such as Native Americans [17]. This larger sample may provide additional insight into how food insecure households rely on HFP to minimize or lessen their food insecurity in new ways. Second, our analysis is specifically focused on the COVID-19 pandemic, an unprecedented time in recent history, in which unemployment and job loss, as well as food supply chain disruptions were widespread, triggering levels of food insecurity not seen in decades. Indeed, given that cost savings is often a motivation for HFP [3, 23, 24], such financial and lifestyle disruptions were likely an important component of HFP motivation and increased intensity. Finally, our study asked about a suite of HFP strategies, while other studies have typically focused on a single strategy such as gardening or fishing. This may be especially important when interpreting the results, since a larger percentage of food insecure households as compared to food secure households were engaging in non-gardening activities, which may have different potential impacts on food security. Hunting, fishing, and foraging for example, may not actually secure food in the same ways that gardening or backyard livestock could more reliably, at least during the time period in which our survey was conducted (e.g. summer before major hunting seasons).

Second, our results demonstrate clear links between HFP and diet quality outcomes, especially for current fruit and vegetable intake among respondents using AHFP, gardening and canning, and for current red meat intake among hunters and those with backyard livestock. These results confirm previous research findings that gardening is correlated with increased fruit and vegetable intake (e.g. [25, 29, 31]. While other research on hunting and fishing has not linked these behaviors to higher intake of red meat, prior research among a Native American population found that hunting, fishing and foraging increased the diversity and quality of diets [17]. While red meat intake is linked to various adverse health outcomes (e.g.[58–61]), not all red meats have the same nutritional profile. Wild meat and game that could be acquired through hunting may provide important micronutrients and protein [62, 63], providing important dietary quality benefits.

These findings may have important long-term health implications, especially the finding that nearly one in four respondents was eating less fruits and vegetables during the COVID-19 pandemic than before. Increased fruit and vegetable intake is associated with reduced risk of cardiovascular disease, certain cancers, and all-cause mortality [64], yet even pre-pandemic, most Americans did not meet the national recommendations for fruit and vegetable intake [65]. Our finding of reduced intake are similar to those from studies conducted recently in France (Constant et al. 2020) and the United Arab Emirates [37] finding lower fruit and vegetable intake during COVID-19 associated lockdowns. Respondents using AHFP were on average eating ½ cup more each of fruits and vegetables daily; higher fruit and vegetable intake is associated with reduced risk of cardiovascular disease, cancer and mortality [64]. Furthermore, since previous research suggests that gardening is also associated with improved nutritional knowledge [33, 34], and long-term beneficial changes in eating habits [24, 35], the significant uptick in gardening and other HFP strategies during the pandemic may have future impacts on diet quality and health not yet realized. Future research should continue to monitor these potential changes, including their link to health outcomes more specifically.

There are many opportunities to expand this work with future research and address potential limitations of the current study. One limitation of this study is a lack of understanding about the amount of food generated through HFP activities. Future research could more clearly explore how different quantities of HFP affect food security and diet quality outcomes by asking what percent of food intake is coming from HFP, or whether HFP activities, especially hunting, fishing and foraging, result in food procurement. Further, in some of our diet quality metrics, we combined red and processed meat, which may have different nutritional profiles, especially if wild meat is part of a diet. These should be more carefully separated in future studies. Secondly, this work demonstrates outcomes during a global pandemic, when many people’s daily lives were significantly changed.

People potentially had new motivations for pursuing HFP activities that could be related to food security, but also may be unrelated (e.g. hobbies, time in nature, cultural trends). Long-term potential diet and food security costs and benefits from HFP will likely accrue over many years. Therefore, it is critical to assess whether the new and increased intensity of HFP is sustained in the future. Such sustained efforts would also potentially have important impacts on conservation through increased demand in hunting, fishing and foraging that should be adequately considered. As well, long-term increased engagement in HFP activities may require increased resources for people pursuing these activities through educational efforts and opportunities for low-cost infrastructure, especially since gardening can have significant up-front costs [26]. Finally, given the social distanced nature of COVID-19, this research utilized an online survey to capture an understanding of this issue, but this research would certainly benefit from additional qualitative and quantitative data analysis. Interviews and focus groups could contextualize the results and better understand the motivations and challenges of HFP activities, which can provide important information for future education and resource allocation. Future studies would benefit from a longitudinal or interventional design that support the examination of causality.

## Conclusion

This study documented the extent of a range of HFP activities among a statewide sample in the US and assessed associations between HFP and food security and dietary outcomes. The results demonstrate that HFP activities significantly increased during the first five months of the COVID-19 pandemic, and were especially prominent among food insecure households. The results also document clear relationships between HFP activities and dietary outcomes, including higher fruit and vegetable intake, and possibly increased diet diversity, which may have important health benefits long-term. Taken together, the results suggest that HFP activities are an important, and potentially increasingly important, way in which many people engage in the food system and the natural environment, with potential implications for both conservation and nutrition and health outcomes. As such, additional research should more fully aim to understand these relationships over time, and in greater depth, especially in the continuation and aftermath of the COVID-19 pandemic. As well, additional collaborations within the conservation sector may be important to assess the long-term impact of increased levels of HFP that may affect forests, waterways, and species. Heightened engagement in HFP may necessitate expanded education and outreach efforts to provide resources for HFP that is productive and sustainable.

## Data Availability

The survey instruments are available at Harvard Dataverse: https://dataverse.harvard.edu/dataverse/foodaccessandcoronavirus

## Declarations

### Ethics approval and consent to participate

Institutional Review Board approval was obtained from the University of Vermont under protocol 00000873. Consent was obtained from all participants prior to data collection.

### Consent for publication

Not Applicable

### Availability of data and materials

The survey instrument materials used for this current study are available at Harvard Dataverse at: https://dataverse.harvard.edu/dataverse/foodaccessandcoronavirus. The datasets used and/or analysed during the current study are available from the corresponding author on reasonable request.

### Authors’ contributions

MTN conceived and designed the work, analyzed the data, and wrote and revised the manuscript. KBW conceived and designed the work, wrote and revised the manuscript. EHB and FB conceived and designed the work, and revised the manuscript.

## Acknowledgements

We thank Peter Callas for statistical advice for this analysis. This research is conducted as part of the National Food Access and COVID Research Team (NFACT). NFACT is a national collaboration of researchers committed to rigorous, comparative, and timely food access research during the time of COVID-19. We do this through collaborative, open access research that prioritizes communication to key decision-makers while building our scientific understanding of food system behaviors and policies. <u>To learn more visit www.nfactresearch.org.</u>

## References

1. Callen J. New Household Pulse Survey Shows Concern Over Food Security, Loss of Income. US Census Bureau. 2020. https://www.census.gov/library/stories/2020/05/new-household-pulse-survey-shows-concern-over-food-security-loss-of-income.html. Accessed 2 Feb 2021.

2. Taylor JR, Lovell ST. Urban home gardens in the Global North: A mixed methods study of ethnic and migrant home gardens in Chicago, IL. Renew Agric Food Syst. 2015;30:22–32. doi:DOI: 10.1017/S1742170514000180.

3. Nova P, Pinto E, Chaves B, Silva M. Urban organic community gardening to promote environmental sustainability practices and increase fruit, vegetables and organic food consumption. Gac Sanit. 2020;34:4–9. doi:https://doi.org/10.1016/j.gaceta.2018.09.001.

4. Friedman A. The strange allure of pioneer living. The Atlantic. 2018. https://www.theatlantic.com/magazine/archive/2018/11/shaye-elliott-homesteading/570796/.

5. Elkhoraibi C, Blatchford RA, Pitesky ME, Mench JA. Backyard chickens in the United States: A survey of flock owners. Poult Sci. 2014;93:2920–31. doi:https://doi.org/10.3382/ps.2014-04154.

6. US Department of Interior. 2016 National survey of fishing, hunting and wildlife-associated recreation. 2016. https://www.fws.gov/wsfrprograms/subpages/nationalsurvey/nat_survey2016.pdf.

7. Zhang X, Miller CA. Associations between socioeconomic status and hunting license sales among census tracts in Cook County, Illinois. Hum Dimens Wildl. 2019;24:148–58. doi:10.1080/10871209.2018.1558466.

8. D’Amelio J. Victory Gardens for the war against COVID-19. CBS News. 2020. https://www.cbsnews.com/news/victory-gardens-for-the-war-against-covid-19/.

9. Rao T. Food supply anxiety brings back Victory Gardens. The New York Times. 2020. https://www.nytimes.com/2020/03/25/dining/%0Avictory-gardens-coronavirus.html%0A.

10. Flesher J, Nichols AL. Hunting licenses soar as virus-weary Americans head outdoors. Associated Press. 2020. https://apnews.com/article/hunting-licenses-soar-us-outdoors-38cb1118ff3f2844e94dc3e8f3d274a6.

11. Rathke L. Some gardeners in a pickle over scarce canning supplies. Associated Press. 2020. https://apnews.com/article/west-virginia-vermont-virus-outbreak-gardening-archive-867645663bf3eb60bd8d1df161f7020b.

12. Sachdeva S, Emery MR, Hurley PT. Depiction of Wild Food Foraging Practices in the Media: Impact of the Great Recession. Soc Nat Resour. 2018;31:977–93. doi:10.1080/08941920.2018.1450914.

13. Lingeman RR. Don’t you know there’s a war on? The American home front 1941-1945. New York: G.P. Putnam’s Sons; 1970.

14. Niles MT, Bertmann F, Belarmino EH, Wentworth T, Biehl E, Neff RA. The Early Food Insecurity Impacts of COVID-19. Nutrients. 2020;12:2096.

15. Wolfson JA, Leung CW. Food Insecurity and COVID-19: Disparities in Early Effects for US Adults. Nutrients. 2020;12.

16. Corrigan MP. Growing what you eat: Developing community gardens in Baltimore, Maryland. Appl Geogr. 2011;31:1232–41. doi:https://doi.org/10.1016/j.apgeog.2011.01.017.

17. Smith E, Ahmed S, Dupuis V, Crane MR, Eggers M, Pierre M, et al. Contribution of Wild Foods to Diet, Food Security, and Cultural Values Amidst Climate Change. J Agric Food Syst Community Dev. 2019;9 B SE-Indigenous Food Sovereignty Peer-Reviewed Papers. doi:10.5304/jafscd.2019.09B.011.

18. Huet C, Rosol R, Egeland GM. The Prevalence of Food Insecurity Is High and the Diet Quality Poor in Inuit Communities. J Nutr. 2012;142:541–7. doi:10.3945/jn.111.149278.

19. Cooke SJ, Twardek WM, Lennox RJ, Zolderdo AJ, Bower SD, Gutowsky LFG, et al. The nexus of fun and nutrition: Recreational fishing is also about food. Fish Fish. 2018;19:201–24. doi:https://doi.org/10.1111/faf.12246.

20. Toth JF, Brown RB. Racial and gender meanings of why people participate in recreational fishing. Leis Sci. 1997;19:129–46. doi:10.1080/01490409709512244.

21. Hunt KM, Floyd MF, Ditton RB. African-American and Anglo Anglers’ Attitudes toward the Catch-Related Aspects of Fishing. Hum Dimens Wildl. 2007;12:227–39. doi:10.1080/10871200701442825.

22. Darby K, Hinton T, Torre J. The Motivations and Needs of Rural, Low-Income Household Food Gardeners. J Agric Food Syst Community Dev. 2020;9 2 SE-Open Call Papers. doi:10.5304/jafscd.2020.092.002.

23. Algert SJ, Baameur A, Renvall MJ. Vegetable Output and Cost Savings of Community Gardens in San Jose, California. J Acad Nutr Diet. 2014;114:1072–6. doi:10.1016/j.jand.2014.02.030.

24. Gray L, Guzman P, Glowa KM, Drevno AG. Can home gardens scale up into movements for social change? The role of home gardens in providing food security and community change in San Jose, California. Local Environ. 2014;19:187–203. doi:10.1080/13549839.2013.792048.

25. Algert S, Diekmann L, Renvall M, Gray L. Community and home gardens increase vegetable intake and food security of residents in San Jose, California. Calif Agric. 2016;70:77–82.

26. Csortan G, Ward J, Roetman P. Productivity, resource efficiency and financial savings: An investigation of the current capabilities and potential of South Australian home food gardens. PLoS One. 2020;15:e0230232. https://doi.org/10.1371/journal.pone.0230232.

27. Strunk C, Richardson M. Cultivating belonging: refugees, urban gardens, and placemaking in the Midwest, U.S.A. Soc Cult Geogr. 2019;20:826–48. doi:10.1080/14649365.2017.1386323.

28. Hoover C, Ostertag S, Hornby C, Parker C, Hansen-Craik, K., Loseto L, Pearce T. The continued important of hunting for future Inuit food security. Solut J. 2016;7:40–50.

29. Alaimo K, Packnett E, Miles RA, Kruger DJ. Fruit and Vegetable Intake among Urban Community Gardeners. J Nutr Educ Behav. 2008;40:94–101. doi:https://doi.org/10.1016/j.jneb.2006.12.003.

30. Litt JS, Soobader M-J, Turbin MS, Hale JW, Buchenau M, Marshall JA. The Influence of Social Involvement, Neighborhood Aesthetics, and Community Garden Participation on Fruit and Vegetable Consumption. Am J Public Health. 2011;101:1466–73. doi:10.2105/AJPH.2010.300111.

31. Barnidge EK, Hipp PR, Estlund A, Duggan K, Barnhart KJ, Brownson RC. Association between community garden participation and fruit and vegetable consumption in rural Missouri. Int J Behav Nutr Phys Act. 2013;10:128. doi:10.1186/1479-5868-10-128.

32. Stark PB, Miller D, Carlson TJ, de Vasquez KR. Open-source food: Nutrition, toxicology, and availability of wild edible greens in the East Bay. PLoS One. 2019;14:e0202450. https://doi.org/10.1371/journal.pone.0202450.

33. Machida D, Kushida O. The Influence of Food Production Experience on Dietary Knowledge, Awareness, Behaviors, and Health among Japanese: A Systematic Review. International Journal of Environmental Research and Public Health. 2020;17.

34. Palar K, Lemus Hufstedler E, Hernandez K, Chang A, Ferguson L, Lozano R, et al. Nutrition and Health Improvements After Participation in an Urban Home Garden Program. J Nutr Educ Behav. 2019;51:1037–46. doi:10.1016/j.jneb.2019.06.028.

35. Kortright R, Wakefield S. Edible backyards: a qualitative study of household food growing and its contributions to food security. Agric Human Values. 2011;28:39–53. doi:10.1007/s10460-009-9254-1.

36. Savoie-Roskos MR, Wengreen H, Durward C. Increasing Fruit and Vegetable Intake among Children and Youth through Gardening-Based Interventions: A Systematic Review. J Acad Nutr Diet. 2017;117:240–50. doi:10.1016/j.jand.2016.10.014.

37. Cheikh Ismail L, Osaili TM, Mohamad MN, Al Marzouqi A, Jarrar AH, Abu Jamous DO, et al. Eating Habits and Lifestyle during COVID-19 Lockdown in the United Arab Emirates: A Cross-Sectional Study. Nutrients. 2020;12.

38. Constant A, Conserve DF, Gallopel-Morvan K, Raude J. Socio-Cognitive Factors Associated With Lifestyle Changes in Response to the COVID-19 Epidemic in the General Population: Results From a Cross-Sectional Study in France. Frontiers in Psychology. 2020;11:2407. https://www.frontiersin.org/article/10.3389/fpsyg.2020.579460.

39. Belarmino, E.H., Bertmann F, Wentworth T, Biehl E, Neff R, Niles MT. The impact of COVID-19 on the local food system: Early findings from Vermont. 2020. https://scholarworks.uvm.edu/calsfac/23.

40. Chenarides L, Grebitus C, Lusk JL, Printezis I. Who practices urban agriculture? An empirical analysis of participation before and during the COVID-19 pandemic. Agribusiness. 2020;n/a n/a. doi:https://doi.org/10.1002/agr.21675.

41. Sofo A, Sofo A. Converting Home Spaces into Food Gardens at the Time of Covid-19 Quarantine: all the Benefits of Plants in this Difficult and Unprecedented Period. Hum Ecol. 2020;48:131–9. doi:10.1007/s10745-020-00147-3.

42. Lal R. Home gardening and urban agriculture for advancing food and nutritional security in response to the COVID-19 pandemic. Food Secur. 2020;12:871–6. doi:10.1007/s12571-020-01058-3.

43. Piontak JR, Schulman MD. Food Insecurity in Rural America. Contexts. 2014;13:75–7. doi:10.1177/1536504214545766.

44. Niles MT., Neff R., Biehl E, Bertmann, Farryl; Morgan, Emily H.; Wentworth T. Food Access and Security During Coronavirus Survey-Version 1.0. 2020.

45. Niles MT, Belarmino EH, Bertmann F, Biehl E, Acciai F, Josephson A, et al. Food insecurity during COVID-19: A multi-state research collaborative. medRxiv. 2020;:2020.12.01.20242024. doi:10.1101/2020.12.01.20242024.

46. Niles MT., Neff R, Biehl E, Bertmann F, Belarmino, Emily H.; Acciai F, Ohri-Vachaspati P. Food Access and Food Security During COVID-19 Survey-Version 2.1. 2020. https://doi.org/10.7910/DVN/4KY9XZ.

47. Peterson RA. A Meta-Analysis of Cronbach’s Coefficient Alpha. J Consum Res. 1994;21:381–91. http://www.jstor.org/stable/2489828.

48. Nunnally JC. Pscyhometric Theory. 2nd edition. New York: McGraw-Hill; 1978.

49. Bureau UC. ACS Demographic and housing estimates for Vermont. 2019. https://data.census.gov/cedsci/table?g=0400000US50&tid=ACSDP5Y2019.DP05.

50. Vermont Department of Health. Vermont population estimates and census data. 2020. https://www.healthvermont.gov/health-statistics-vital-records/vital-records-population-data/vermont-population-estimates. Accessed 31 Dec 2020.

51. USDA Economic Research Service. U.S. household food security survey module: six-item short form. 2012. https://www.ers.usda.gov/topics/food-nutrition-assistance/food-security-in-the-us/survey-tools/#six.

52. Yaroch AL, Tooze J, Thompson FE, Blanck HM, Thompson OM, Colón-Ramos U, et al. Evaluation of Three Short Dietary Instruments to Assess Fruit and Vegetable Intake: The National Cancer Institute’s Food Attitudes and Behaviors Survey. J Acad Nutr Diet. 2012;112:1570–7. doi:10.1016/j.jand.2012.06.002.

53. Stuart EA. Matching Methods for Causal Inference: A Review and a Look Forward. Stat Sci. 2010;25:1– 21. doi:10.1214/09-STS313.

54. Hill JL, Reiter JP, Zanutto EL. A comparison of experimental and observational data analyses. Appl Bayesian Model Causal Inference from Incomplete-Data Perspect An Essent Journey with Donald Rubin’s Stat Fam. 2004;:49–60.

55. Rubin DB, Thomas N. Matching using estimated propensity scores: relating theory to practice. Biometrics. 1996;:249–64.

56. Rubin DB. Using multivariate matched sampling and regression adjustment to control bias in observational studies. J Am Stat Assoc. 1979;74:318–28.

57. Zhao Z. Using matching to estimate treatment effects: Data requirements, matching metrics, and Monte Carlo evidence. Rev Econ Stat. 2004;86:91–107.

58. Cordain L, Watkins BA, Florant GL, Kelher M, Rogers L, Li Y. Fatty acid analysis of wild ruminant tissues: evolutionary implications for reducing diet-related chronic disease. Eur J Clin Nutr. 2002;56:181–91.

59. Lippi G, Mattiuzzi C, Cervellin G. Meat consumption and cancer risk: a critical review of published meta-analyses. Crit Rev Oncol Hematol. 2016;97:1–14. doi:https://doi.org/10.1016/j.critrevonc.2015.11.008.

60. Wang X, Lin X, Ouyang YY, Liu J, Zhao G, Pan A, et al. Red and processed meat consumption and mortality: dose–response meta-analysis of prospective cohort studies. Public Health Nutr. 2016;19:893–905.

61. Pan A, Sun Q, Bernstein AM, Schulze MB, Manson JE, Stampfer MJ, et al. Red meat consumption and mortality: results from 2 prospective cohort studies. Arch Intern Med. 2012;172:555–63.

62. Mann N. Dietary lean red meat and human evolution. Eur J Nutr. 2000;39:71–9.

63. Strazdina V, Jemeljanovs A, Šterna V. Nutrition value of wild animal meat. In: Proceedings of the Latvian Academy of Sciences. Section B. Natural, Exact, and Applied Sciences. Sciendo; 2013. p. 373–7.

64. Aune D, Giovannucci E, Boffetta P, Fadnes LT, Keum N, Norat T, et al. Fruit and vegetable intake and the risk of cardiovascular disease, total cancer and all-cause mortality—a systematic review and dose-response meta-analysis of prospective studies. Int J Epidemiol. 2017;46:1029–56. doi:10.1093/ije/dyw319.

65. Moore L V, Dodd KW, Thompson FE, Grimm KA, Kim SA, Scanlon KS. Using Behavioral Risk Factor Surveillance System Data to Estimate the Percentage of the Population Meeting US Department of Agriculture Food Patterns Fruit and Vegetable Intake Recommendations. Am J Epidemiol. 2015;181:979–88. doi:10.1093/aje/kwu461.

