## Supplementary Materials for "Home Food Procurement Impacts Food Security and Diet Quality during COVID-19"

Supplementary Table 1. Results of a logit model predicting food security with demographic controls.

| Variable Name | Odds Ratio | Std. Err. | p= | 95% Confidence Interval | |
|---|---|---|---|---|---|
| Female | 1.052 | 0.261 | 0.840 | 0.646 | 1.710 |
| Children in HH | 0.842 | 0.253 | 0.567 | 0.467 | 1.518 |
| Over 55 | 2.518 | 0.674 | 0.001 | 1.490 | 4.256 |
| BIPOC/Hispanic | 1.606 | 0.663 | 0.251 | 0.715 | 3.605 |
| Negative Job Change | 0.474 | 0.106 | 0.001 | 0.306 | 0.733 |
| Less50k | 0.134 | 0.031 | 0.000 | 0.085 | 0.211 |
| HH Size | 0.857 | 0.089 | 0.139 | 0.699 | 1.051 |

Supplementary Table 2. Food insecurity by disaggregated race and ethnicity.

| | Food Insecurity Rate | | | |
|---|---|---|---|---|
| | For Demographic Group | For Outside Demographic Group | Total in Demographic Group | p= (chi2 test) |
| Asian | 25.0% | 29.1% | 4 | 0.858 |
| Black | 50.0% | 28.7% | 8 | 0.188 |
| Native American | 20.0% | 29.1% | 5 | 0.655 |
| Multiple Race | 33.3% | 28.9% | 21 | 0.066 |
| White | 28.7% | 34.2% | 544 | 0.467 |
| BIPOC/hispanic | 36.2% | 28.4% | 47 | 0.261 |
| Hispanic | 50.0% | 28.4% | 16 | 0.061 |

Supplementary Table 3. Logit model predicting gardening activity since COVID-19 by demographic controls.

| Variable | Odds Ratio | Std. Error | p= | 95% Confidence Interval | |
|---|---|---|---|---|---|
| Female | 0.905 | 0.176 | 0.606 | 0.618 | 1.324 |
| Children in HH | 1.341 | 0.360 | 0.274 | 0.793 | 2.269 |
| Over 55 | 1.351 | 0.291 | 0.162 | 0.886 | 2.060 |
| BIPOC/Hispanic | 0.884 | 0.295 | 0.711 | 0.459 | 1.700 |
| Negative Job Change | 1.425 | 0.263 | 0.055 | 0.993 | 2.047 |
| Less $50K | 0.632 | 0.118 | 0.014 | 0.438 | 0.912 |
| HH Size | 0.867 | 0.080 | 0.124 | 0.723 | 1.040 |

Supplementary Table 4. Logit model predicting fishing activity since COVID-19 by demographic controls.

| Variable | Odds Ratio | Std. Error | p= | 95% Confidence Interval | |
|---|---|---|---|---|---|
| Female | 0.732 | 0.220 | 0.299 | 0.407 | 1.318 |
| Children in HH | 1.265 | 0.484 | 0.539 | 0.598 | 2.677 |
| Over 55 | 0.501 | 0.177 | 0.051 | 0.250 | 1.003 |
| BIPOC/Hispanic | 1.095 | 0.517 | 0.848 | 0.434 | 2.763 |
| Negative Job Change | 1.550 | 0.449 | 0.131 | 0.878 | 2.735 |
| Less $50K | 0.718 | 0.211 | 0.258 | 0.404 | 1.276 |
| HH Size | 0.939 | 0.125 | 0.634 | 0.723 | 1.218 |

Supplementary Table 5. Logit model predicting foraging activity since COVID-19 by demographic controls.

| Variable | Odds Ratio | Std. Error | p= | 95% Confidence Interval | |
|---|---|---|---|---|---|
| Female | 1.207 | 0.397 | 0.567 | 0.633 | 2.302 |
| Children in HH | 1.260 | 0.535 | 0.586 | 0.549 | 2.895 |
| Over 55 | 1.846 | 0.682 | 0.097 | 0.894 | 3.809 |
| BIPOC/Hispanic | 1.162 | 0.594 | 0.769 | 0.427 | 3.162 |
| Negative Job Change | 2.130 | 0.652 | 0.014 | 1.169 | 3.881 |
| Less $50K | 1.463 | 0.438 | 0.204 | 0.813 | 2.630 |
| HH Size | 1.119 | 0.158 | 0.427 | 0.848 | 1.477 |

Supplementary Table 6. Logit model predicting hunting activity since COVID-19 by demographic controls.

| Variable | Odds Ratio | Std. Error | p= | 95% Confidence Interval | |
|---|---|---|---|---|---|
| Female | 0.458 | 0.169 | 0.034 | 0.222 | 0.942 |
| Children in HH | 2.263 | 1.105 | 0.094 | 0.870 | 5.890 |
| Over 55 | 0.594 | 0.277 | 0.265 | 0.238 | 1.483 |
| BIPOC/Hispanic | 1.529 | 0.812 | 0.424 | 0.540 | 4.328 |
| Negative Job Change | 1.642 | 0.614 | 0.185 | 0.789 | 3.416 |
| Less $50K | 0.900 | 0.333 | 0.777 | 0.436 | 1.859 |
| HH Size | 0.836 | 0.144 | 0.299 | 0.597 | 1.172 |

Supplementary Table 7. Logit model predicting backyard livestock activity since COVID-19 by demographic controls.

| Variable | Odds Ratio | Std. Error | p= | 95% Confidence Interval | |
|---|---|---|---|---|---|
| Female | 0.721 | 0.276 | 0.392 | 0.340 | 1.526 |
| Children in HH | 1.248 | 0.560 | 0.621 | 0.518 | 3.005 |
| Over 55 | 0.157 | 0.091 | 0.001 | 0.050 | 0.491 |
| BIPOC/Hispanic | 1.404 | 0.740 | 0.519 | 0.500 | 3.944 |
| Negative Job Change | 1.487 | 0.551 | 0.285 | 0.719 | 3.075 |
| Less $50K | 0.741 | 0.271 | 0.412 | 0.362 | 1.518 |
| HH Size | 0.962 | 0.149 | 0.800 | 0.710 | 1.303 |

Supplementary Table 8. Logit model predicting canning activity since COVID-19 by demographic controls.

| Variable | Odds Ratio | Std. Error | p= | 95% Confidence Interval | |
|---|---|---|---|---|---|
| Female | 0.920 | 0.198 | 0.698 | 0.604 | 1.402 |
| Children in HH | 1.342 | 0.395 | 0.318 | 0.754 | 2.388 |
| Over 55 | 1.317 | 0.321 | 0.257 | 0.818 | 2.122 |
| BIPOC/Hispanic | 1.417 | 0.485 | 0.309 | 0.724 | 2.773 |
| Negative Job Change | 1.454 | 0.297 | 0.067 | 0.973 | 2.171 |
| Less $50K | 0.703 | 0.147 | 0.091 | 0.467 | 1.058 |
| HH Size | 0.963 | 0.097 | 0.704 | 0.790 | 1.172 |

Supplementary Table 9. Percent of Respondents by Food Security Status Engaging in HFP activities. P values determined through chi-square tests.

| Activity | Food Secure | Food Insecure | p= |
|---|---|---|---|
| Any home food procurement | 39.2% | 47.3% | 0.076 |
| More HFP since COVID | 44.4% | 66.2% | 0.002 |
| Gardens Since | 34.3% | 35.5% | 0.782 |
| Fishing Since | 7.7% | 15.4% | 0.005 |
| Foraging Since | 7.0% | 14.8% | 0.003 |
| Hunting Since | 3.9% | 11.8% | 0.000 |
| Livestock Since | 4.3% | 10.1% | 0.008 |
| Canning Since | 20.5% | 29.6% | 0.019 |
| Gardens More | 38.8% | 58.9% | 0.005 |
| Fishing More | 28.3% | 51.1% | 0.025 |
| Foraging More | 34.3% | 59.4% | 0.040 |
| Hunting More | 14.3% | 47.2% | 0.003 |
| Livestock More | 47.6% | 60.0% | 0.401 |
| Canning More | 53.1% | 63.3% | 0.136 |

Supplementary Table 10. Matching results examining current fruit intake, with various treatment variables. Each row indicate a separate matching result.

| Variable | Coefficient | Robust Std. Error | p= | 95% Confidence Interval | | Model n= |
|---|---|---|---|---|---|---|
| Any HFP | 0.386 | 0.116 | 0.001 | 0.158 | 0.613 | 565 |
| HFP More | 0.010 | 0.18 | 0.954 | -0.342 | 0.363 | 240 |
| Garden Since | 0.329 | 0.121 | 0.006 | 0.092 | 0.565 | 571 |
| Foraging Since | 0.036 | 0.259 | 0.891 | -0.472 | 0.544 | 571 |
| Canning Since | 0.240 | 0.133 | 0.071 | -0.021 | 0.501 | 571 |
| Gardens More | -0.038 | 0.187 | 0.838 | -0.405 | 0.328 | 227 |
| Foraging More | 0.276 | 0.439 | 0.528 | -0.583 | 1.136 | 67 |
| Canning More | -0.066 | 0.243 | 0.786 | -0.543 | 0.411 | 155 |

Supplementary Table 11. Matching results examining current vegetable intake, with various treatment variables. Each row indicates a separate matching result.

| Variable | Coefficient | Robust Std. Error | p= | 95% Confidence Interval | | Model n= |
|---|---|---|---|---|---|---|
| Any HFP | 0.526 | 0.118 | 0.000 | 0.295 | 0.756 | 565 |
| HFP More | 0.019 | 0.162 | 0.908 | -0.299 | 0.336 | 240 |
| Garden Since | 0.541 | 0.119 | 0.000 | 0.307 | 0.775 | 571 |
| Foraging Since | 0.360 | 0.229 | 0.115 | -0.088 | 0.808 | 571 |
| Canning Since | 0.511 | 0.135 | 0.000 | 0.246 | 0.776 | 571 |
| Gardens More | -0.002 | 0.181 | 0.990 | -0.357 | 0.353 | 227 |
| Foraging More | 0.183 | 0.381 | 0.632 | -0.564 | 0.929 | 67 |
| Canning More | -0.291 | 0.240 | 0.225 | -0.762 | 0.180 | 155 |

Supplementary Table 12. Matching results examining current red meat intake, with various treatment variables. Each row indicates a separate matching result.

| Variable | Coefficient | Robust Std. Error | p= | 95% Confidence Interval | | Model n= |
|---|---|---|---|---|---|---|
| Any HFP | 0.145 | 0.157 | 0.355 | -162.000 | 0.451 | 565 |
| HFP More | -0.099 | 0.234 | 0.671 | -0.557 | 0.358 | 240 |
| Fishing Since | 0.210 | 0.209 | 0.315 | -0.200 | 0.620 | 571 |
| Hunting Since | 0.527 | 0.246 | 0.032 | 0.045 | 1.009 | 571 |
| Livestock Since | 0.794 | 0.243 | 0.001 | 0.316 | 1.271 | 571 |
| Fishing More | -0.418 | 0.348 | 0.230 | -1.100 | 0.264 | 93 |
| Hunting More | -0.348 | 0.409 | 0.395 | -1.150 | 0.454 | 71 |
| Livestock More | -0.271 | 0.468 | 0.562 | -1.187 | 0.645 | 49 |

Supplementary Table 13. Matching results examining current processed meat intake, with various treatment variables. Each row indicates a separate matching result.

| Variable | Coefficient | Robust Std. Error | p= | 95% Confidence Interval | | Model n= |
|---|---|---|---|---|---|---|
| Any HFP | -0.196 | 9.156 | 0.208 | -0.501 | 0.109 | 565 |
| HFP More | 0.124 | 0.243 | 0.610 | -0.352 | 0.600 | 240 |
| Fishing Since | 0.134 | 0.251 | 0.594 | -0.359 | 0.627 | 571 |
| Hunting Since | -0.292 | 0.402 | 0.468 | -1.080 | 0.496 | 571 |
| Livestock Since | 0.188 | 0.339 | 0.580 | -0.477 | 0.852 | 571 |
| Fishing More | 0.438 | 0.428 | 0.307 | -0.401 | 1.276 | 93 |
| Hunting More | -0.102 | 0.501 | 0.839 | -1.084 | 0.880 | 71 |
| Livestock More | 0.510 | 0.531 | 0.336 | -0.530 | 1.550 | 49 |

Supplementary Table 14. Matching results examining change in fruit and vegetable intake since COVID-19, with various treatment variables. Each row indicates a separate matching result.

| Variable | Coefficient | Robust Std. Error | p= | 95% Confidence Interval | | Model n= |
|---|---|---|---|---|---|---|
| Any HFP | 0.066 | 0.049 | 0.183 | -0.031 | 0.162 | 565 |
| HFP More | -0.004 | 0.079 | 0.954 | -0.159 | 0.159 | 240 |
| Garden Since | 0.048 | 0.055 | 0.383 | -0.06 | 0.156 | 571 |
| Foraging Since | 0.027 | 0.096 | 0.779 | -0.161 | 0.215 | 571 |
| Canning Since | 0.087 | 0.056 | 0.12 | -0.023 | 0.197 | 571 |
| Gardens More | -0.027 | 0.084 | 0.74 | -0.192 | 0.138 | 227 |
| Foraging More | -0.143 | 0.166 | 0.387 | -0.467 | 0.181 | 67 |
| Canning More | -0.06 | 0.109 | 0.583 | -0.275 | 0.154 | 155 |

Supplementary Table 15. Matching results examining change in red and processed meat intake since COVID-19, with various treatment variables. Each row indicates a separate matching result.

| Variable | Coefficient | Robust Std. Error | p= | 95% Confidence Interval | | Model n= |
|---|---|---|---|---|---|---|
| Any HFP | -0.025 | 0.049 | 0.617 | -0.121 | 0.072 | 564 |
| HFP More | -0.057 | 0.076 | 0.455 | -0.206 | 0.092 | 239 |
| Fishing Since | 0.091 | 0.089 | 0.303 | -0.083 | 0.265 | 570 |
| Hunting Since | 0.083 | 0.116 | 0.473 | -0.144 | 0.31 | 570 |
| Livestock Since | 0.036 | 0.134 | 0.789 | -0.227 | 0.298 | 570 |
| Fishing More | -0.046 | 0.177 | 0.792 | -0.393 | 0.3 | 93 |
| Hunting More | -0.502 | 0.201 | 0.021 | -0.895 | 0.108 | 71 |
| Livestock More | -0.079 | 0.223 | 0.722 | -0.517 | 0.357 | 49 |
